# Comparing genome-wide significant and chemosensory variants as instruments for dietary patterns in Mendelian randomisation

**DOI:** 10.64898/2026.03.26.26349354

**Authors:** Phoebe SC Hui, Brooke L Devlin, David M Evans, Liang-Dar Hwang

**Author notes:** Corresponding author; 306 Carmody Rd, St Lucia QLD 4067, Australia.

## Abstract

**Background:** Diet is a modifiable risk factor for cardiometabolic disease, yet establishing causality remains challenging. Mendelian randomisation (MR) leverages genetic variants as instrumental variables (IVs) to enable causal inference.

**Method:** Using two-sample MR, we assessed the causal effects of four principal component-derived dietary patterns (DPs) – Unhealthy, Healthy, Meat-based, Pescatarian – on twelve cardiometabolic outcomes: body mass index, coronary artery disease, high-density lipoprotein cholesterol, low-density lipoprotein cholesterol, total cholesterol, triglycerides, systolic blood pressure, diastolic blood pressure, type 2 diabetes, fasting glucose and insulin, and glycated haemoglobin. Two sets of IVs were employed: conventional genome-wide significant variants associated with each DP, rigorously filtered for pleiotropy and directionality; and biologically informed variants in chemosensory receptor genes, given the role of taste and smell perception in shaping food choices.

**Results:** Using conventional IVs, the Pescatarian DP reduced fasting insulin (β_IVW_ = -0.10 pmolL^-1^ per SD increase in Pescatarian DP score, 95% Confidence interval [CI] [-0.15, - 0.04]; *P* = 1.19×10^-3^), which survived multiple sensitivity analyses. Associations between the Unhealthy DP and elevated blood pressure and glycated haemoglobin were likely undermined by heterogeneity and pleiotropy, with insufficient IVs for robust sensitivity testing. Chemosensory receptors yielded null findings, reflecting insufficient power.

**Conclusion:** Rigorously filtered conventional IVs supported the causal nature of well-established diet-disease relationships, demonstrating MR’s utility in strengthening causal inference in nutritional epidemiology. Chemosensory IVs demonstrated limited utility for DPs, likely reflecting the heterogeneous and complex sensory profiles of overall diets. Future efforts should consider using guideline-based dietary scores to facilitate translation of findings.

**Key messages:**

- We used Mendelian randomisation to evaluate the causal effects of dietary patterns on cardiometabolic health and explored whether variants in chemosensory receptor genes could improve instrument specificity given their mechanistic link to food choice.
- Compared with variants selected from dietary genome-wide association studies (GWAS) solely based on statistical significance, chemosensory receptor variants showed fewer associations with confounding pathways but explained insufficient variance in composite dietary patterns to support well-powered MR analyses.
- Using rigorously selected genome-wide significant instruments, a Pescatarian-like pattern was associated with lower fasting insulin, but effects across other glycaemic markers and disease endpoints were limited, suggesting that diet alone only has modest direct effects on health when isolated from non-dietary factors in MR.

## Background

Diet is a major risk factor for cardiometabolic diseases, yet causal inference remains challenging. Observational epidemiology is susceptible to unmeasured confounding and reverse causation, while long-term dietary randomised controlled trials (RCTs) face practical and ethical challenges [1]. Mendelian randomisation (MR) offers an alternative by using genetic variants as instrumental variables (IVs) to proxy modifiable risk factors, such as diet [1–3]. As alleles are randomly allocated at meiosis, MR can be conceptualised as nature’s RCT, enabling causal inference from observational data. For valid causal estimates, three core assumptions must hold: (i) the genetic variants are robustly associated with the exposure; (ii) no confounders are associated with both the variant and outcome; and (iii) the variant only (potentially) affects the outcome through the exposure, with no alternative pathways [4].

Most diet-related MR studies have examined single foods or nutrients [5, 6]. However, dietary exposures are typically compositional, where changes in one food imply changes in another food in the opposite direction [6]. Genome-wide significant variants for specific dietary components may therefore capture correlated intakes rather than just the intended exposure [6], introducing horizontal pleiotropy. Dietary patterns (DPs), such as those derived using principal component analysis (PCA), can capture food combinations and substitutions that occur in real-world eating behaviours [7], and recent genome-wide association studies (GWAS) have identified genetic variants associated with these patterns [8–10].

However, GWAS-derived DP instruments raise methodological concerns. DP GWAS often identified variants in gene regions such as *FTO*, *APOE*, and *FGF21* [9–11], which have broad metabolic effects and may influence cardiometabolic outcomes independently of DP (**Figure 1, orange arrows**). These variants may also influence DPs indirectly, particularly in older adults who may have modified their diets following diagnosis or risk assessment [8]. Consequently, associations observed in GWAS may reflect reverse causation from health status to DP rather than a genetic predisposition to dietary preferences, leading to misspecification of the exposure if these variants are used as MR instruments (**Figure 1, purple arrows**). Bidirectional MR has demonstrated that being diagnosed with coronary heart disease could shift people towards healthier DPs [9, 10]. Approaches such as Steiger filtering of instruments [12] or phenome-wide association studies (PheWAS) can help identify variants with incorrect directionality or potentially pleiotropic associations [5]. Prioritising variants with functional or biological relevance to the exposure trait over those chosen purely on statistical grounds may also reduce the risk of both horizontal pleiotropy and reverse causation [3, 5, 13].

Variants related to chemosensory perception offer one such biologically grounded option for dietary exposures [14]. Chemosensory receptor variants influence taste, smell, and chemesthetic perception through receptor-mediated detection of food compounds, creating individual variability in flavour sensitivity that shape food preference and choices [15, 16]. Receptor variants have been linked to specific dietary behaviours, including lower intake of vegetables [17], added salt [18], and red wine [18] among individuals with heightened bitter taste sensitivity, ultimately shaping broader DPs. This biological specificity means that chemosensory-associated variants are less likely to influence cardiometabolic outcomes through non-dietary pathways, reducing the risk of horizontal pleiotropy and reverse causation (**Figure 1, bottom panel**). Chemosensory receptor variants with consistent effects on food preferences across young and late adulthood have been used as IVs for specific food items in MR studies of cardiometabolic health [14].

This study applies MR to examine the causal effects of DPs on cardiometabolic outcomes while addressing these methodological challenges. In particular, cardiometabolic outcomes were selected as their relationships with diet are well established and can provide a benchmark for validating the use of MR and biologically informed instruments for complex dietary exposures. Two complementary sets of instruments were used: (1) genome-wide significant (*P* < 5×10^-8^) variants from DP GWAS as primary IVs, with comprehensive IV filtering and sensitivity analyses to detect and control for potential confounding and horizontal pleiotropy; and (2) chemosensory receptor variants as biologically informed instruments less likely to influence outcomes through non-dietary pathways.

## Methods

### Study design

This study followed a two-sample MR framework, using summary-level data from existing GWAS to estimate the causal effects of DPs on cardiometabolic outcomes. The overall workflow is illustrated in **Figure 2**. Reporting followed the STROBE-MR checklist [19].

### Data Sources

Details for all GWAS are summarised in **Table S1**. GWAS summary statistics for DPs were obtained from three UK Biobank studies, where dietary intake was assessed using a touchscreen questionnaire with 29 diet-related items [8–10]. Four distinct DPs were derived using PCA on questionnaire responses, with methodological differences across studies outlined in **Table 1**.

Cardiometabolic outcomes were selected based on well-documented associations with diet, including body mass index (BMI) [20], coronary artery disease (CAD) [21], high-density lipoprotein cholesterol (HDL-C), low-density lipoprotein cholesterol (LDL-C), total cholesterol (TC), total triglycerides (TG) [22], systolic blood pressure (SBP), diastolic blood pressure (SBP) [23], type 2 diabetes (T2D) [24], haemoglobin A1c (HbA1c), fasting glucose (FG), and fasting insulin (FI) [25]. CAD and T2D were analysed as case-control outcomes, with effect estimates representing log odds ratios [21, 24]. Lipid traits were adjusted for cholesterol-lowering medication use where applicable [22], while glycaemic markers were measured in people without type 1 diabetes (T1D) or T2D [25].

Analyses were restricted to European ancestry to minimise potential bias from population stratification. Where possible, outcome GWAS summary statistics were obtained from meta-analyses excluding UK Biobank participants, although overlap remained for CAD, SBP, DBP and T2D (40%, 45%, 45% and 10%, respectively).

### Instrument selection and quality control

‘Conventional’ IVs were the independent, genome-wide significant variants reported in the source GWAS for each DP [8–10]. ‘Receptor’ IVs were sourced from 1214 nonsynonymous single-nucleotide polymorphisms (SNPs) within non-pseudo taste and olfactory receptor genes with minor allele frequency (MAF) ≥ 0.01 in the UK Biobank identified by Hwang et al. [14], which were then screened for associations with each DP (FDR-corrected *P* < 0.05; F-statistics > 10) and clumped at 500 kb and r^2^ < 0.01. SNPs within 1 Mb of the major histocompatibility complex (MHC) region were excluded as extensive linkage disequilibrium (LD) in this region may compromise the exclusion-restriction assumption.

All candidate IVs underwent a two-stage PheWAS using ieugwasr v1.0.2 [26], scanning for associations across OpenGWAS (accessed 2 April 2025) [27, 28] at *P* < 1×10^-5^, approximating a Bonferroni-corrected threshold (0.05/4358 traits = 1.15×10^-5^) previously used by Cole et al. [9] based on the number of traits in the UK Biobank [27, 28]. This correction is inherently conservative as a proportion of the GWAS queried involved overlapping individuals and correlated traits. Stage 1 (*a priori* sources of bias) flagged associations between the candidate IVs with early-life traits (traits with temporal precedence over adult DP formation, i.e., childhood anthropometry [age <18 years], birth outcomes) and adult behavioural and socioeconomic traits commonly considered in observational nutritional epidemiology: socioeconomic status (SES) (including income, housing tenure, number of vehicles, Townsend deprivation index), educational attainment, alcohol consumption, physical activity, and sedentary time **(Table S2**). Flagged SNPs were either: (1) removed from univariable MR; or (2) adjusted for using multivariable MR (MVMR) models by incorporating these traits as additional exposures [29, 30]. Stage 2 (exploratory) reviewed remaining PheWAS associations for additional pleiotropic pathways. Adult anthropometric and clinical traits were not treated as sources of bias, as these may lie downstream of DPs. Instead, directionality was addressed using Steiger filtering [12], retaining only SNPs explaining more variance in the DP than in any of the 12 outcomes.

Palindromic IVs were aligned using allele frequencies, where variants with allele frequencies between 0.40 and 0.60 were treated as ambiguous and omitted from all MR analyses. IVs absent from an outcome dataset were replaced with proxies (r^2^ > 0.8 in Europeans) identified via ieugwasr for outcomes available in OpenGWAS [27, 28], LDproxy [31], followed by HaploReg v4.2 if still unavailable [32]. Manually identified proxies are listed in **Table S4**. All remaining variants constitute the post-quality control (post-QC) IV set.

For MVMR, additional exposures were selected to avoid collinearity by choosing, within each trait domain (e.g., household income, Townsend deprivation index, and housing tenure all representing SES), the UK Biobank-derived GWAS with greatest overlap between its instruments and DP IVs. Five traits were included alongside each DP: educational attainment, household income, physical activity, smoking status [33], and comparative body size at age 10 [34] (**Table S1**).

Instrument strength in MVMR was assessed via the Sanderson-Windmeijer conditional F[statistics (MVMR R package v0.4) [35]. Phenotypic covariances among exposures in each MVMR model were estimated using UK Biobank individual-level data (*n* = 275 950 unrelated participants of European ancestry). DP scores were reconstructed following the original GWAS methods [8–10]. For the Unhealthy DP, defined using genetic correlations between intakes of individual food items, PCA was performed on a genetic correlation matrix reconstructed from the published pairwise genetic correlation estimates [8]. Non-dietary traits were also coded and adjusted per their respective GWAS protocols [33, 34].

### 2.4 Genetic correlation analysis

Bivariate Linkage Disequilibrium Score (LDSC) regression (v1.0.1) [36, 37] was used to estimate genetic correlations (r_g_) among the four DPs and the cardiometabolic outcomes. SNPs were restricted to the HapMap3 reference panel and excluded for MAF < 0.01, reference allele mismatches, strand ambiguity, or (where available) imputation INFO < 0.9. LD scores were calculated using the 1000 Genomes Phase 3 European reference panel.

The effective number of independent tests was determined using matSpD [38] (https://research.qut.edu.au/sgel/software/matspd-local-version/) based on the genetic correlation matrices. The four DPs represented three independent exposures. Outcomes were grouped by consortium and analysed in blocks to account for sample overlap, yielding an effective number of 11 independent outcomes. Accordingly, the Bonferroni-corrected significance threshold for the present study was α = 0.05/(3×11) = 0.00152.

### Mendelian randomisation

All MR analyses were conducted in R v4.4.2 using TwoSampleMR (v0.6.19) [12, 28], ieugwasr (v1.0.4) [26], and MRPRESSO (v1.0) [39]. Causal effects were estimated using the inverse variance-weighted (IVW) method. IV validity was assessed using Cochran’s Q statistic for heterogeneity in causal effect size estimates and the MR Egger intercept for directional pleiotropy [40].

### Sensitivity analyses

MR Egger [41, 40] and the weighted median estimator [42] were used to obtain causal effect estimates that are more robust to invalid IVs and to confirm the direction of effect indicated by IVW. Where heterogeneity was detected, MR-PRESSO was used to estimate causal effects with outliers removed [39].

### Power calculations

Power was estimated using the equations of Brion et al. [43] to determine the minimum detectable effect sizes at 80% power for α = 0.05 and α = 0.00152 (Bonferroni correction). For binary outcomes, alternative equations provided through the online power calculator (https://shiny.cnsgenomics.com/mRnd/) were used. As these equations were developed for one-sample MR, the power estimates (**Table S5**) are for approximation only.

### Ethics approval

This study primarily used publicly available GWAS summary statistics. UK Biobank individual-level data were accessed under application 53641 for estimating phenotypic covariances between exposures in MVMR. The UK Biobank holds ethical approval from the North West Multi-Centre Research Committee (21/NW/0157).

## Results

### SNP associations with dietary patterns

From the original DP GWAS, we identified 16, 60, 29, and 63 Conventional IVs for the Unhealthy, Healthy, Meat-based, and Pescatarian DPs, respectively. Two Pescatarian DP SNPs were removed due to proximity to the MHC region. After removing SNPs associated with behavioural, socioeconomic or early-life traits (*P* < 1×10^-5^) and those failing Steiger filtering, post-QC Conventional IVs were two, 22, 12, and 30 respectively for the Unhealthy, Healthy, Meat-based, and Pescatarian DPs, with no overlapping IVs between DPs.

For Receptor IVs, initial counts were one, eight, one, and seven for the Unhealthy, Healthy, Meat-based, and Pescatarian DPs. No Receptor IVs were associated with the behavioural, socioeconomic, or early-life traits. Instead, four Receptor IVs were excluded due to proximity to the MHC region, and one Pescatarian IV failed Steiger filtering for SBP and DBP. Removing these SNPs resulted in seven post-QC Receptor IVs for the Healthy DP, four for the Pescatarian DP, and none for the Unhealthy and Meat-based DPs remaining (**Table S6**).

Associations of each IV with its respective DP and with each outcome, alongside variance explained, Steiger filtering, and PheWAS-based QC results, are reported in **Tables S7** and **S8**, respectively, for Conventional and Receptor SNPs.

### Results from main analysis using Conventional instruments

Using conventional IVs that passed QC filters (see Methods), four causal associations were observed at the Bonferroni-corrected threshold (*P* < 1.52×10^-3^). Each standard deviation (SD) increase in the Unhealthy DP score was associated with higher SBP (β_IVW_ = 2.54 mmHg per SD, 95% CI [1.53, 3.56]; *P* = 8.64×10^-7^), DBP (β_IVW_ = 1.07 mmHg per SD, 95% CI [0.44, 1.70]; *P* = 9.03×10^-4^), and HbA1c (β_IVW_ = 0.13% per SD, 95% CI [0.06, 0.20]; *P* = 2.54×10^-4^), while each SD increase in Pescatarian DP scores was associated with reduced FI (β_IVW_= - 0.10 pmolL^-1^ per SD, 95% CI [-0.15, -0.04]; *P* = 1.19×10^-3^). MR effect estimates for all DP-outcome pairs are shown in **Figure 3** and **Table S9**.

No heterogeneity was detected for these four causal associations (Cochran’s Q *P* > 0.05), although heterogeneity was observed across DP-outcome pairs that yielded non-significant associations with IVW (**Table S10**). For the Unhealthy DP, despite only two instruments, heterogeneity was detected in five outcomes, with rs12568918 showing inconsistent directions of effect across cardiometabolic traits (**Table S6**). This suggests potential horizontal pleiotropy through pathways independent of DP, despite the SNP passing Steiger filtering for directionality. The remaining IV, rs4410790, is strongly associated with caffeine consumption [44] and explained more variance in coffee intake than in the Unhealthy DP (R^2^ for coffee = 46.6%; R^2^ for Unhealthy DP = 7.1%) [8], suggesting that findings may reflect the effects of caffeine rather than broader DPs. Directional pleiotropy could not be assessed for the Unhealthy DP associations as fewer than three IVs were available, while the Pescatarian DP-FI association showed no evidence of directional pleiotropy (Egger intercept *P* > 0.05). Complete heterogeneity and pleiotropy statistics are provided in, respectively, **Table S10** and **S11**.

### Results from sensitivity and MVMR analyses using Conventional instruments

Findings from sensitivity analyses using the weighted median estimator, MR-Egger, MR-PRESSO, and MVMR are presented along with the primary IVW estimates in **Table S9**.

For the Unhealthy DP, the weighted median estimator, MR-Egger, and MR-PRESSO could not be performed due to the limited number of IVs. MVMR showed effect directions consistent with the primary IVW results, but with wide confidence intervals overlapping the null and evidence for weak instrument bias (conditional F_Unhealthy_ = 2.96; **Table S12**).

For the Pescatarian DP-FI association, weighted median and MR-Egger estimates were directionally consistent with IVW, but confidence intervals overlapped with the null. MR-PRESSO did not detect any outlier IVs. MVMR replicated this association at nominal significance (*P* = 3.02 ×10^-2^) with conditional F-statistics indicating weak instruments (F_Pescatarian_ = 2.90; for selected traits = range 1.84-9.67; see **Table S12**). As such, MVMR estimates were interpreted cautiously and only used to assess consistency of effect direction with the primary IVW analysis.

### Results from main analysis using Receptor instruments

MR estimates using Receptors IVs are provided in **Table S13**. Upon MHC exclusions, no IVs remained for the Unhealthy and Meat-based DPs. For the Healthy and Pescatarian DPs, all estimates overlapped with the null (**Figure 4**), with heterogeneity in the causal estimates and directional pleiotropy detected (*P* < 0.05) for some associations (**Tables S10 & S11**).

### Results from sensitivity analysis using Receptor instruments

All weighted median and MR Egger estimates overlapped with the null, except for a suggestive association between the Healthy DP and reduced SBP (weighted median *P* = 4.05×10^-2^). MR-PRESSO either did not detect any outliers or that outlier-corrected estimates overlapped with the null.

## Discussion

This study used MR to investigate the causal effects of DPs on cardiometabolic outcomes, emphasising rigorous IV selection and attempting to compare biologically informed against conventional, significance-based IV selection strategies. Overall, evidence for independent causal effects of broad DPs on cardiometabolic outcomes was mixed. The Pescatarian DP showed a consistent effect on reducing FI across sensitivity analyses. The apparent effects of the Unhealthy DP on blood pressure and HbA1c should be interpreted cautiously; after filtering out 58 of 60 IVs with heterogeneous associations across cardiometabolic traits or caffeine metabolism, the remaining two IVs precluded pleiotropy-robust sensitivity analyses. Sample overlap between the DP and blood pressure GWAS may further bias these estimates towards the observed phenotypic associations [45, 46]. Chemosensory receptor IVs yielded null findings across DP-outcome pairs, likely reflecting insufficient statistical power.

The protective effect of a genetically predicted Pescatarian DP on FI aligns with observational [47, 48] and experimental [49] evidence for Mediterranean-style diets, which emphasise oily fish, vegetables, fruits, and minimal processed meat. Fish consumption in particular may improve insulin sensitivity via omega-3 fatty acids reducing lipid-induced impairment of insulin signalling [47], while dietary fibre has been associated with improved fasting insulin in RCTs, potentially through the gut microbiota [50]. However, the Pescatarian DP showed no effects on FG, HbA1c, and T2D risk, possibly reflecting that dietary components influence distinct physiological pathways. More broadly, null findings for downstream outcomes (T2D) may suggest that DP alone, when isolated from other risk factors through MR, may be insufficient to produce clinically detectable changes in disease risk, consistent with diet functioning as one causal risk factor within a multifactorial disease process.

Null findings for chemosensory receptor IVs likely reflect both statistical and biological limitations. Biologically informed IVs may offer higher specificity but typically explain less variance in the exposure compared to a larger collection of genome-wide significant SNPs [3]. The effects of chemosensory variants appear to be stronger for individual food items than for composite DPs [14], with their signals diluted when aggregated across multiple foods.

This reduced explained variance, coupled with fewer IVs, meant that the chemosensory approach had low statistical power in detecting small to moderate causal effects if they exist [43]. “Winner’s curse” [51] may further bias effect estimates towards the null. Future research should prioritise identifying additional biologically relevant variants with stronger effects on DPs, or developing statistical methods that aggregate pathway-specific IVs for individual food items (e.g., chemosensory receptor variants for specific food items, lactase persistence for dairy intake) to model DPs, thereby retaining biological specificity.

A broader challenge lies in translating MR findings using PCA-derived DPs into practical recommendations, as they reflect relative intake combinations rather than absolute quantities. Guideline-based indices with absolute intake targets, such as the Healthy Eating Index (HEI) [52], may offer clearer translational value, as the scoring criteria use real-world food group units, allowing individuals or practitioners to identify which components fall short of defined targets and act accordingly. Unlike PCA-derived patterns, where loadings and scale shift across study populations, fixed scoring frameworks help retain interpretability across different contexts. Wider adoption of such indices in dietary MR requires consistent availability of GWAS summary statistics for specific *a priori* patterns, while PCA remains valuable for studying associations between existing DPs and health outcomes in real-world populations.

## Conclusion

This study provides limited evidence for causal effects of DPs on cardiometabolic health, with the most consistent finding being a protective effect between a Pescatarian DP and FI. Rigorous IV selection and multivariable models were applied to address pleiotropy, and chemosensory receptor variants were explored as biologically informed instruments, though insufficient variance explained in composite DPs limited statistical power. Future work should prioritise GWAS of guideline-based diet quality scores and methods that aggregate pathway-specific variants across food items to model overall DPs.

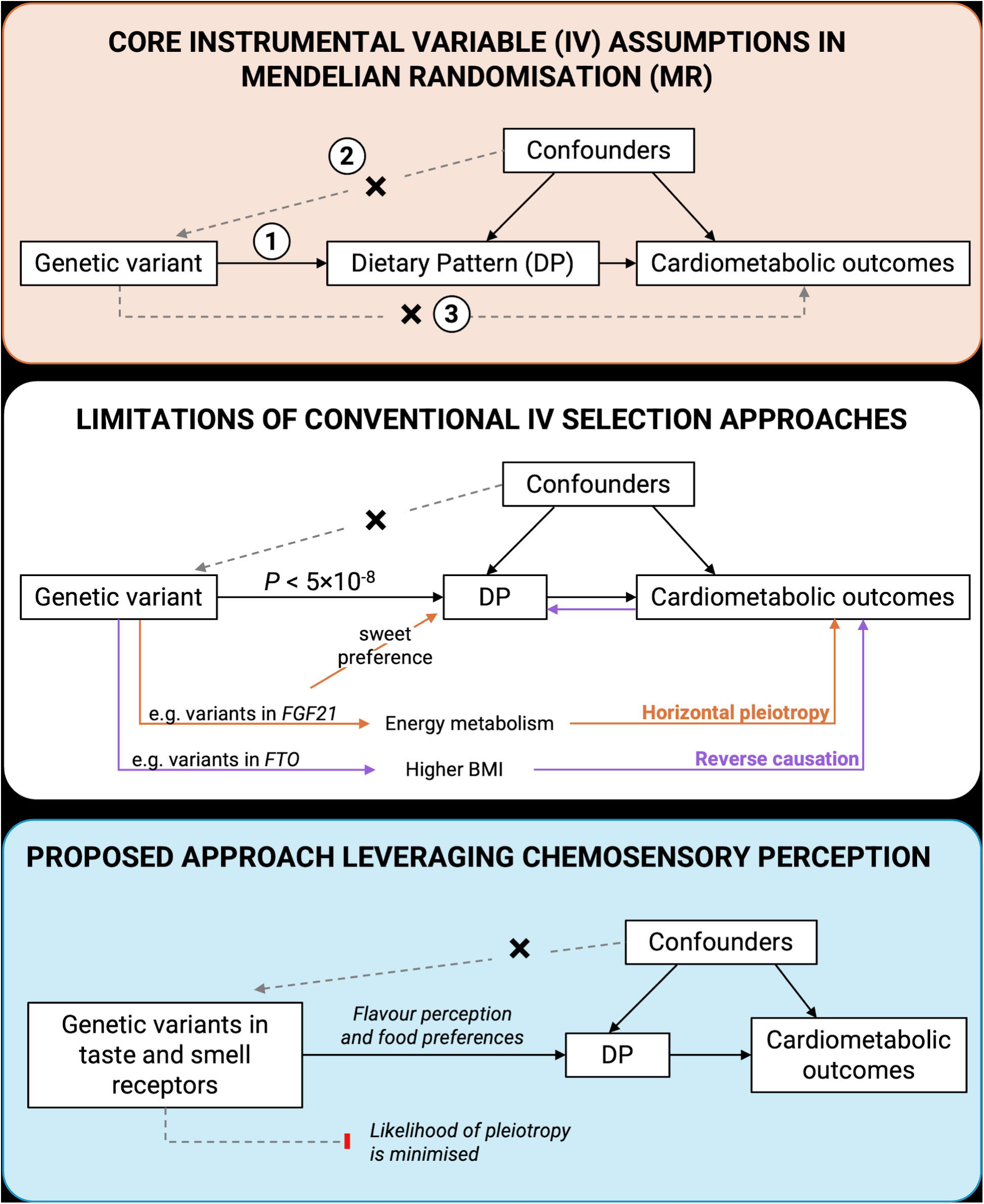

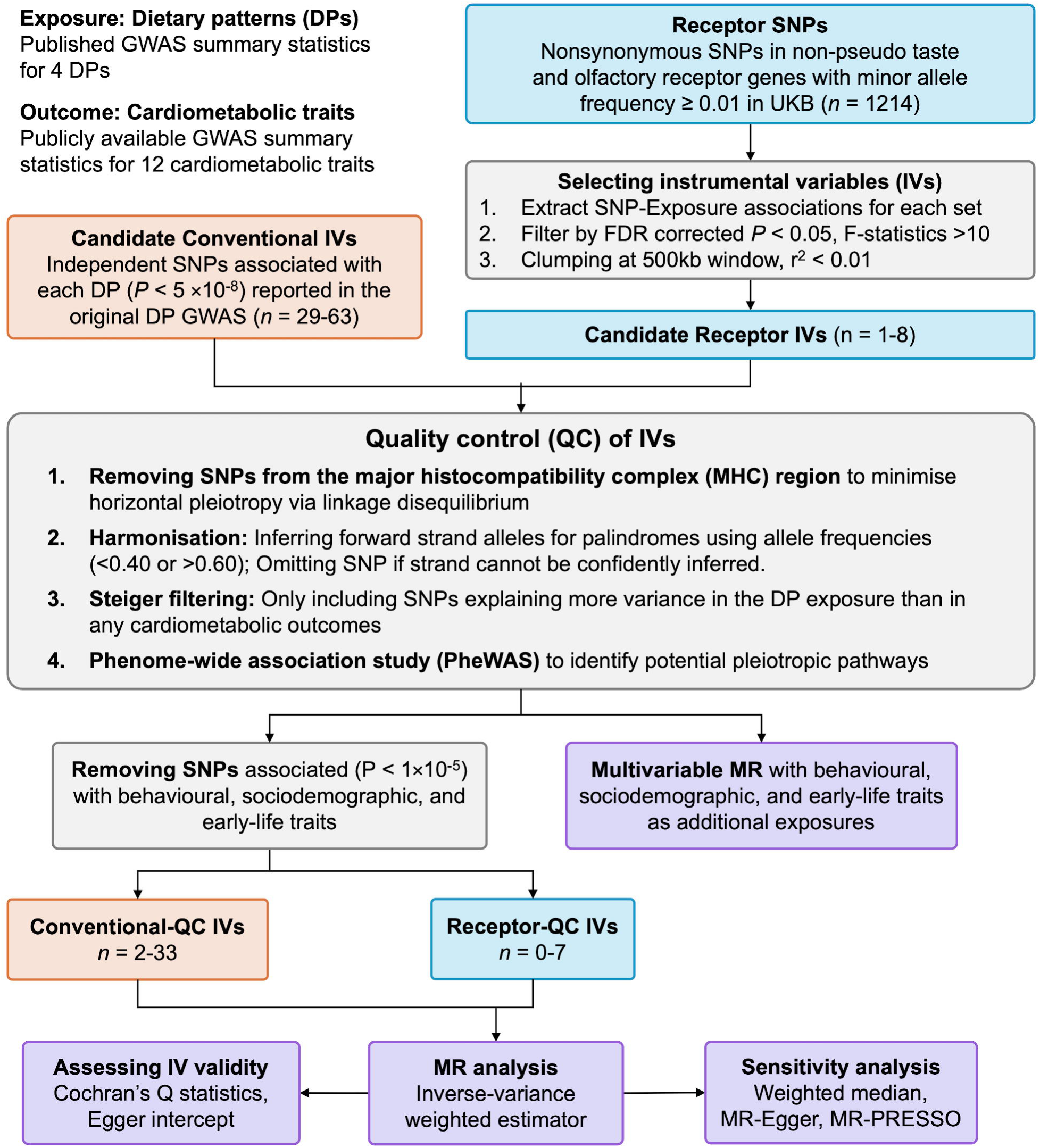

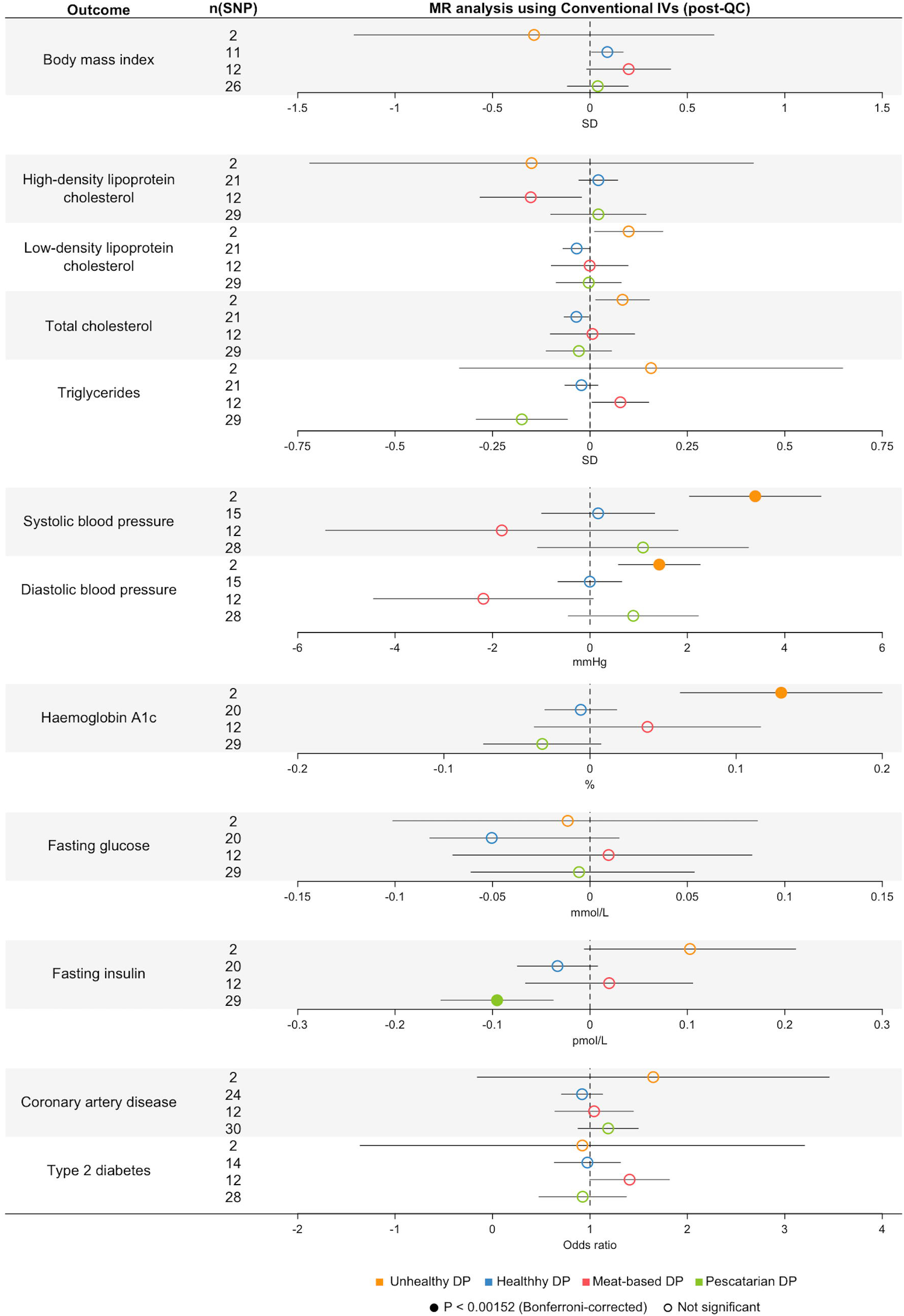

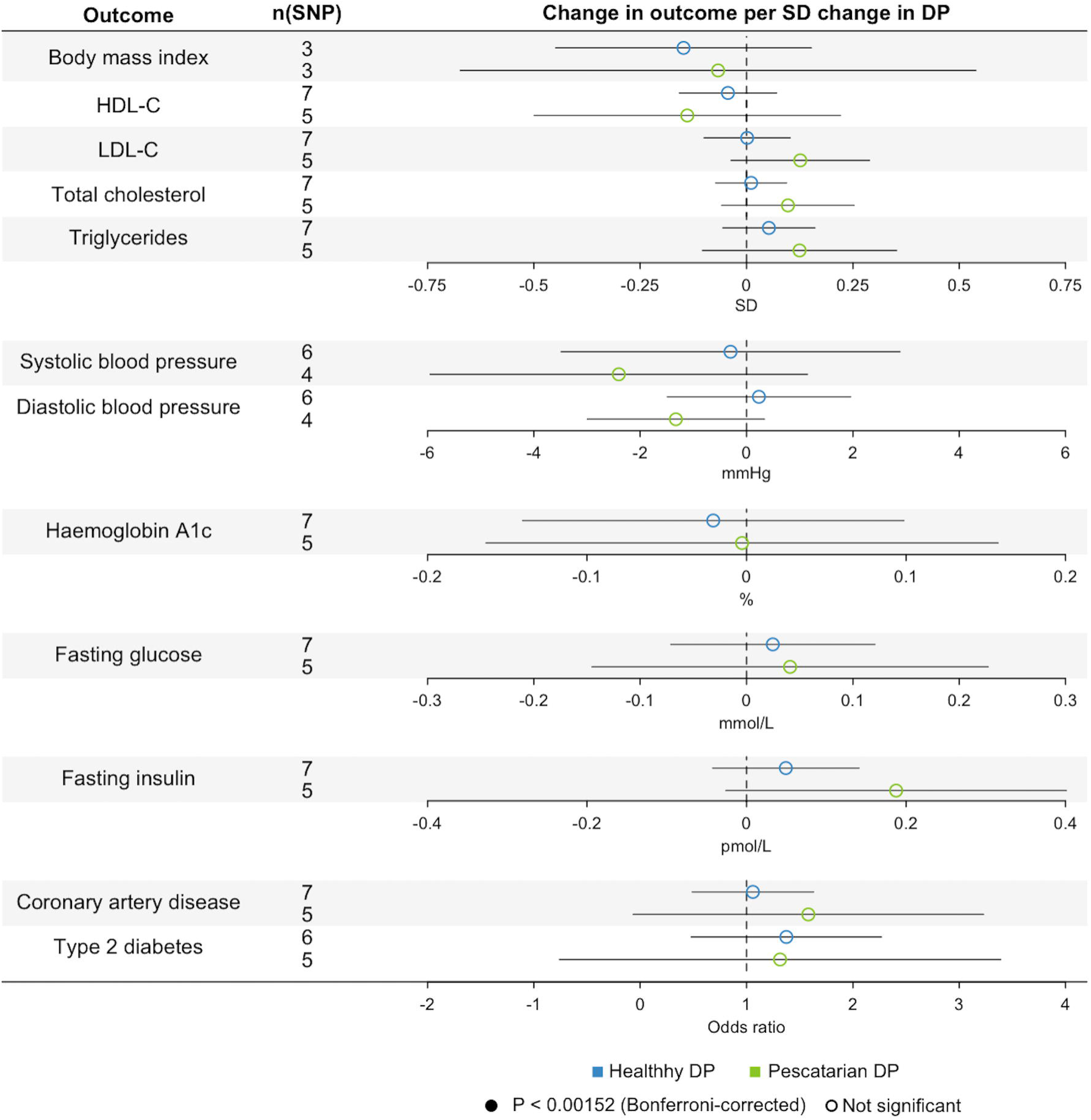

## Data Availability

All data produced in the present study are available upon reasonable request to the authors

## Data availability

GWAS summary statistics of the Meat-based and Pescatarian DP were obtained from the authors of Niarchou et al. All other GWAS summary statistics used in this study are publicly available from the GWAS Catalog (https://www.ebi.ac.uk/gwas/) and the OpenGWAS website (https://gwas.mrcieu.ac.uk). UK Biobank individual-level data are available to approved researchers from the UK Biobank (http://www.ukbiobank.ac.uk/).

## Acknowledgement

This research was conducted using UK Biobank Resource under Application Number 53641. We used publicly available genome-wide association study (GWAS) summary statistics from the following sources: dietary exposures (Cole et al., 2020; Niarchou et al., 2020; Pirastu et al., 2022), coronary artery disease from the CARDIoGRAMplusC4D Consortium, body mass index from the GIANT Consortium, blood lipid traits from the Global Lipids Genetics Consortium, glycaemic traits from the MAGIC Consortium, blood pressure (Keaton et al., 2024), and data accessed via the MRC-IEU OpenGWAS project. We gratefully acknowledge the participants and investigators of these studies, including the UK Biobank, whose contributions made this research possible.

## References

[1] Trepanowski JF, Ioannidis JPA. Perspective: Limiting Dependence on Nonrandomized Studies and Improving Randomized Trials in Human Nutrition Research: Why and How. Adv Nutr 2018; 9: 367–377.

[2] Wade KH, Yarmolinsky J, Giovannucci E, et al. Applying Mendelian randomization to appraise causality in relationships between nutrition and cancer. Cancer Causes Control 2022; 33: 631–652.

[3] Haycock PC, Burgess S, Wade KH, et al. Best (but oft-forgotten) practices: the design, analysis, and interpretation of Mendelian randomization studies. Am J Clin Nutr 2016; 103: 965–978.

[4] Sanderson E, Glymour MM, Holmes MV, et al. Mendelian randomization. Nat Rev Methods Primer 2022; 2: 6.

[5] Sutton KJ, Gervis J, Jatoi M, et al. Dietary Intake Mendelian Randomization: Assessment and Development of Methods for Instrument Selection and Robust Inference. Epub ahead of print 27 June 2025. DOI: 10.1101/2025.06.26.25330002.

[6] Merino J, Tobias DK. The unique challenges of studying the genetics of diet and nutrition. Nat Med 2022; 28: 221–222.

[7] McNaughton SA. Dietary patterns. In: Present Knowledge in Nutrition. Elsevier, pp. 235–248.

[8] Pirastu N, McDonnell C, Grzeszkowiak EJ, et al. Using genetic variation to disentangle the complex relationship between food intake and health outcomes. PLOS Genet 2022; 18: e1010162.

[9] Cole JB, Florez JC, Hirschhorn JN. Comprehensive genomic analysis of dietary habits in UK Biobank identifies hundreds of genetic associations. Nat Commun 2020; 11: 1467.

[10] Niarchou M, Byrne EM, Trzaskowski M, et al. Genome-wide association study of dietary intake in the UK biobank study and its associations with schizophrenia and other traits. Transl Psychiatry 2020; 10: 51.

[11] Pirastu N, McDonnell C, Grzeszkowiak EJ, et al. Using genetic variation to disentangle the complex relationship between food intake and health outcomes. PLOS Genet 2022; 18: e1010162.

[12] Hemani G, Tilling K, Davey Smith G. Orienting the causal relationship between imprecisely measured traits using GWAS summary data. PLOS Genet 2017; 13: e1007081.

[13] Burgess S, Woolf B, Mason AM, et al. Addressing the credibility crisis in Mendelian randomization. BMC Med 2024; 22: 374.

[14] Hwang L-D, Lin C, Evans DM, et al. Biologically informed instrument selection for dietary Mendelian randomization using chemosensory receptor variants. Epub ahead of print 6 February 2026. DOI: 10.64898/2026.02.05.26345702.

[15] Feeney EL, McGuinness L, Hayes JE, et al. Genetic variation in sensation affects food liking and intake. Curr Opin Food Sci 2021; 42: 203–214.

[16] Diószegi J, Llanaj E, Ádány R. Genetic Background of Taste Perception, Taste Preferences, and Its Nutritional Implications: A Systematic Review. Front Genet 2019; 10: 1272.

[17] Sandell M, Hoppu U, Mikkilä V, et al. Genetic variation in the hTAS2R38 taste receptor and food consumption among Finnish adults. Genes Nutr 2014; 9: 433.

[18] Brito Nunes C, Lim AW-Y, Dy Q, et al. Phenome-wide investigation of dietary and health outcomes associated with bitter taste receptor gene TAS2R38. Eur J Nutr 2025; 64: 218.

[19] Skrivankova VW, Richmond RC, Woolf BAR, et al. Strengthening the Reporting of Observational Studies in Epidemiology Using Mendelian Randomization: The STROBE-MR Statement. JAMA 2021; 326: 1614.

[20] The LifeLines Cohort Study, The ADIPOGen Consortium, The AGEN-BMI Working Group, et al. Genetic studies of body mass index yield new insights for obesity biology. Nature 2015; 518: 197–206.

[21] Aragam KG, Jiang T, Goel A, et al. Discovery and systematic characterization of risk variants and genes for coronary artery disease in over a million participants. Nat Genet 2022; 54: 1803–1815.

[22] Graham SE, Clarke SL, Wu K-HH, et al. The power of genetic diversity in genome-wide association studies of lipids. Nature 2021; 600: 675–679.

[23] Keaton JM, Kamali Z, Xie T, et al. Genome-wide analysis in over 1 million individuals of European ancestry yields improved polygenic risk scores for blood pressure traits. Nat Genet 2024; 56: 778–791.

[24] Suzuki K, Hatzikotoulas K, Southam L, et al. Genetic drivers of heterogeneity in type 2 diabetes pathophysiology. Nature 2024; 627: 347–357.

[25] Chen J, Spracklen CN, Marenne G, et al. The trans-ancestral genomic architecture of glycemic traits. Nat Genet 2021; 53: 840–860.

[26] Hemani G, Elsworth B, Palmer T, et al. ieugwasr: Interface to the ‘OpenGWAS’ Database API., https://github.com/MRCIEU/ieugwasr (2024).

[27] Elsworth B, Lyon M, Alexander T, et al. The MRC IEU OpenGWAS data infrastructure. bioRxiv. Epub ahead of print 2020. DOI: 10.1101/2020.08.10.244293.

[28] Hemani G, Zheng J, Elsworth B, et al. The MR-Base platform supports systematic causal inference across the human phenome. eLife 2018; 7: e34408.

[29] Sanderson E, Davey Smith G, Windmeijer F, et al. An examination of multivariable Mendelian randomization in the single-sample and two-sample summary data settings. Int J Epidemiol 2019; 48: 713–727.

[30] Burgess S, Thompson SG. Multivariable Mendelian Randomization: The Use of Pleiotropic Genetic Variants to Estimate Causal Effects. Am J Epidemiol 2015; 181: 251–260.

[31] Machiela MJ, Chanock SJ. LDlink: a web-based application for exploring population-specific haplotype structure and linking correlated alleles of possible functional variants. Bioinformatics 2015; 31: 3555–3557.

[32] Ward LD, Kellis M. HaploReg: a resource for exploring chromatin states, conservation, and regulatory motif alterations within sets of genetically linked variants. Nucleic Acids Res 2012; 40: D930–D934.

[33] Elsworth B, Mitchell R, Mitchell R, et al. MRC IEU UK Biobank GWAS pipeline version 2. Epub ahead of print 2019. DOI: 10.5523/BRIS.PNOAT8CXO0U52P6YNFAEKEIGI.

[34] Richardson TG, Sanderson E, Elsworth B, et al. Use of genetic variation to separate the effects of early and later life adiposity on disease risk: mendelian randomisation study. BMJ 2020; m1203.

[35] Sanderson E, Spiller W, Bowden J. Testing and correcting for weak and pleiotropic instruments in two-sample multivariable Mendelian randomization. Stat Med 2021; 40: 5434–5452.

[36] Schizophrenia Working Group of the Psychiatric Genomics Consortium, Bulik-Sullivan BK, Loh P-R, et al. LD Score regression distinguishes confounding from polygenicity in genome-wide association studies. Nat Genet 2015; 47: 291–295.

[37] ReproGen Consortium, Psychiatric Genomics Consortium, Genetic Consortium for Anorexia Nervosa of the Wellcome Trust Case Control Consortium 3, et al. An atlas of genetic correlations across human diseases and traits. Nat Genet 2015; 47: 1236–1241.

[38] Nyholt DR. A Simple Correction for Multiple Testing for Single-Nucleotide Polymorphisms in Linkage Disequilibrium with Each Other. Am J Hum Genet 2004; 74: 765–769.

[39] Verbanck M, Chen C-Y, Neale B, et al. Detection of widespread horizontal pleiotropy in causal relationships inferred from Mendelian randomization between complex traits and diseases. Nat Genet 2018; 50: 693–698.

[40] Burgess S, Thompson SG. Interpreting findings from Mendelian randomization using the MR-Egger method. Eur J Epidemiol 2017; 32: 377–389.

[41] Bowden J, Davey Smith G, Burgess S. Mendelian randomization with invalid instruments: effect estimation and bias detection through Egger regression. Int J Epidemiol 2015; 44: 512–525.

[42] Bowden J, Davey Smith G, Haycock PC, et al. Consistent Estimation in Mendelian Randomization with Some Invalid Instruments Using a Weighted Median Estimator. Genet Epidemiol 2016; 40: 304–314.

[43] Brion M-JA, Shakhbazov K, Visscher PM. Calculating statistical power in Mendelian randomization studies. Int J Epidemiol 2013; 42: 1497–1501.

[44] Cornelis MC, Munafo MR. Mendelian Randomization Studies of Coffee and Caffeine Consumption. Nutrients 2018; 10: 1343.

[45] Lawlor DA. Commentary: Two-sample Mendelian randomization: opportunities and challenges. Int J Epidemiol 2016; 45: 908–915.

[46] Burgess S, Davies NM, Thompson SG. Bias due to participant overlap in two-sample Mendelian randomization. Genet Epidemiol 2016; 40: 597–608.

[47] Vetrani C, Verde L, Colao A, et al. The Mediterranean Diet: Effects on Insulin Resistance and Secretion in Individuals with Overweight or Obesity. Nutrients 2023; 15: 4524.

[48] George ES, Gavrili S, Itsiopoulos C, et al. Poor adherence to the Mediterranean diet is associated with increased likelihood of metabolic syndrome components in children: the Healthy Growth Study. Public Health Nutr 2021; 24: 2823–2833.

[49] Kabthymer RH, Danaher J, De Courten B. Mediterranean diet vs. the Australian guide to healthy eating (AGHE) on body composition and glucose metabolism: A randomized controlled trial. Diabetes Obes Metab 2025; 27: 4689–4698.

[50] Mao T, Huang F, Zhu X, et al. Effects of dietary fiber on glycemic control and insulin sensitivity in patients with type 2 diabetes: A systematic review and meta-analysis. J Funct Foods 2021; 82: 104500.

[51] Jiang T, Gill D, Butterworth AS, et al. An empirical investigation into the impact of winner’s curse on estimates from Mendelian randomization. Int J Epidemiol 2023; 52: 1209–1219.

[52] Reedy J, Lerman JL, Krebs-Smith SM, et al. Evaluation of the Healthy Eating Index-2015. J Acad Nutr Diet 2018; 118: 1622–1633.

