## Supplementary material for "Comparing genome-wide significant and chemosensory variants as instruments for dietary patterns in Mendelian randomisation": Table 1

**Table 1. Description of dietary patterns used as exposures in the present study**

| ***DP*** | ***Description*** | ***DP derivation*** | ***Reference*** |
| --- | --- | --- | --- |
| Overall unhealthy (Unhealthy) | Lower intake of fish, fruit and vegetable + Higher intake of coffee and alcohol + Higher intake of beef, pork, processed meat, lamb, and poultry + More added salt to diet | GWAS was conducted on 29 individual food items, and genetic correlations between the items were estimated via bivariate LD score regression. PCA was applied to the genetic correlation matrix to derive ‘All PC’ (PC1), and GWAS was performed on individual ‘All PC’ scores. | Pirastu et al. [8] |
| Overall healthy (Healthy) | Primarily defined by choosing wholemeal over white bread, along with higher intake of fruit, vegetable, oily fish, and water, and lower intake of processed meat | Diet-related questionnaire responses (35 including nested questions) were converted to 85 food intake quantitative traits (FI-QTs) through ordinal ranking and binary encoding. PCA was then applied to the phenotypic correlation matrix of the 85 FI-QTs to derive PC scores, and GWAS performed on individual PC scores. | Cole et al. [9] |
| Meat-based | High intake of processed meat, poultry, beef, lamb and pork. Orthogonal to the Plant/fish-based diet. | PCA applied on a phenotypic correlation matrix (questionnaire responses for fruit, vegetables, fish, and meat), with the Meat-based DP (‘DC1’) corresponding to PC1. GWAS then performed on individual ‘DC1’ scores. | Niarchou et al. [10] |
| Plant/fish-based (Pescatarian) | High intake of raw and cooked vegetables, fresh and dried fruits, and oily and non-oily fish. Orthogonal to the Meat-based diet. | Corresponds to PC2 (‘DC2’) from the same PCA as above. GWAS performed on individual ‘DC2’ scores. | Niarchou et al. [10] |

*Abbreviations*: DC, dietary component; DP, dietary pattern; GWAS, genome-wide association study; LD, linkage disequilibrium; PC, principal component; PCA, principal component analysis.
